# Predictive Modeling of Heart Failure Readmissions

**DOI:** 10.1101/2025.03.25.25324657

**Authors:** Kevin Felpel, J. W. Awori Hayanga, J Hunter Mehaffey, Christopher Bianco, Marco Caccamo, Vinay Badhwar, George Sokos

## Abstract

**Purpose:** Federal programs to mitigate hospital readmission of patients with heart failure (HF) monetarily encourage hospitals through the use of penalties. The limited performance of predictive models have created potential challenges of implementation and unintended consequences, with criticisms about its unintended consequences and the low performance of its predictive models. We study sought to refine existing predictive models of readmission using heart failure (HF) data from a large multi-payer national dataset.

**Methods:** The Premier healthcare database, a nationally representative all-payor dataset, was utilized to examine over 300 variables from HF patients (2016-2023) including demographics, comorbidities, cardiac diagnoses, provider characteristics, medications, and lab values, defined using diagnosis-related group and ICD-10 codes. Outcomes from patients with primary and secondary HF diagnoses included 30-day all-cause readmissions and 30-day HF-related readmissions. Data were divided into training (60%), validation (20%), and testing (20%) sets. We evaluated logistic regression, random forest, neural networks, modified neural networks, support vector machines, naïve Bayesian decision trees, and XGBoost models, comparing them based on accuracy (AUC), precision, recall, and F-score.

**Results:** Of 722,974 HF patients examined, 12.0% and 11.3% experienced all-cause and HF-related 30-day readmissions, respectively. Mean age was 71 years and 48% were female. A total of 68,649 patients readmitted with a primary HF diagnosis for homogeneity (2021-2023) was thoroughly analyzed using multiple contemporary Bayesian and non-Bayesian models. This subset was 47% female with a mean age of 72 years. The XGBoost model performed best, with an AUC of 0.63 for all-cause and 0.62 for HF-related readmissions. The key predictors of readmissions were age and chronic non-cardiac comorbidities instead of HF-specific factors.

**Conclusion:** Contemporary statistical models applied to nationally representative contemporary real-world data struggle to identify modifiable interventions, suggesting that existing federal programs may penalize without actionable improvements in patient care.

## INTRODUCTION

The rising incidence of heart failure (HF) and its related financial burden has major implications for United States healthcare, with estimated direct costs expected to exceed $70 billion by 2030.^1,2^ The majority of these costs are associated with inpatient care, with approximately one in four patients readmitted within 30 days.^2,3^ In 2007, the Medicare Payment Advisory Commission (MedPAC) approximated that 13.3% of Medicare beneficiaries had preventable heart failure readmissions within 30 days after discharge. This led the Centers for Medicare & Medicaid Services (CMS) to report this metric publicly and preempted the Hospital Readmission Reduction Program (HRRP) under the Affordable Care Act.^4^ The program incentivizes healthcare systems to reduce readmissions through the imposition of a financial penalty of up to 3% for higher-than-expected 30-day readmission rates.

The published all-cause 30-day readmission rate following HF admission is approximately 20%. However, most contemporary predictors of readmission are based on general demographic or diagnosis-related characteristics.^3^ Various prediction models and scores have been derived across different databases, each with unique patient cohorts. Among them include a 2013 study of Medicare beneficiaries in GWTH-HF (Get With The Guidelines – Heart Failure) program, the Readmission After Heart Failure Scale in 2018, and a 2023 study by Kim et al using the Nationwide Readmissions Database.^5–7^ However, national readmission rates remain high despite HRRP financial penalties, implying poor predictors of readmission or the predominance of non-modifiable factors and a prevailing lack of resources to further curb the trend.

We sought to better characterize clinical diagnoses, procedures, and medications in patients admitted with HF and compare those with and without subsequent readmission using a large nationally representative dataset, to identify variables associated with 30-day HF-related, and all-cause readmissions. We hypothesized that non-modifiable patient characteristics unrelated to clinical severity or treatment may be the predominant predictors of readmission.

## METHODS

### Patient Data

The Premier Healthcare Database (PHD) was used to evaluate patients between January 1^st^ 2016 through June 30^th^ 2024 (Table 1). All patients presenting for inpatient care with a primary diagnosis of HF, based on International Classification of Disease (ICD) 10^th^ edition diagnoses were included. The data represent clinical coding, including diagnosis, procedures, and hospital-prescribed medications from approximately 25% of all inpatient hospital admissions throughout the United States (> 1000 hospitals and hospital systems). The database excludes federally funded hospitals (e.g., Veterans Affairs), but otherwise account for a broad national representation based on bed size, geographic region, location (urban/rural) and teaching hospital status. The database contains a date-stamped log of all billed items including medications; laboratory, diagnostic, and therapeutic services; and primary and secondary diagnoses for each patient’s hospitalization. Identifier-linked enrollment files provide demographic and payor information. Detailed service level information for each hospital day is recorded including details on medication and devices received. The West Virginia University Institutional Review Board approved this study with waiver of consent (Protocol #2210660362, 3/23/23).

**Table 1:**
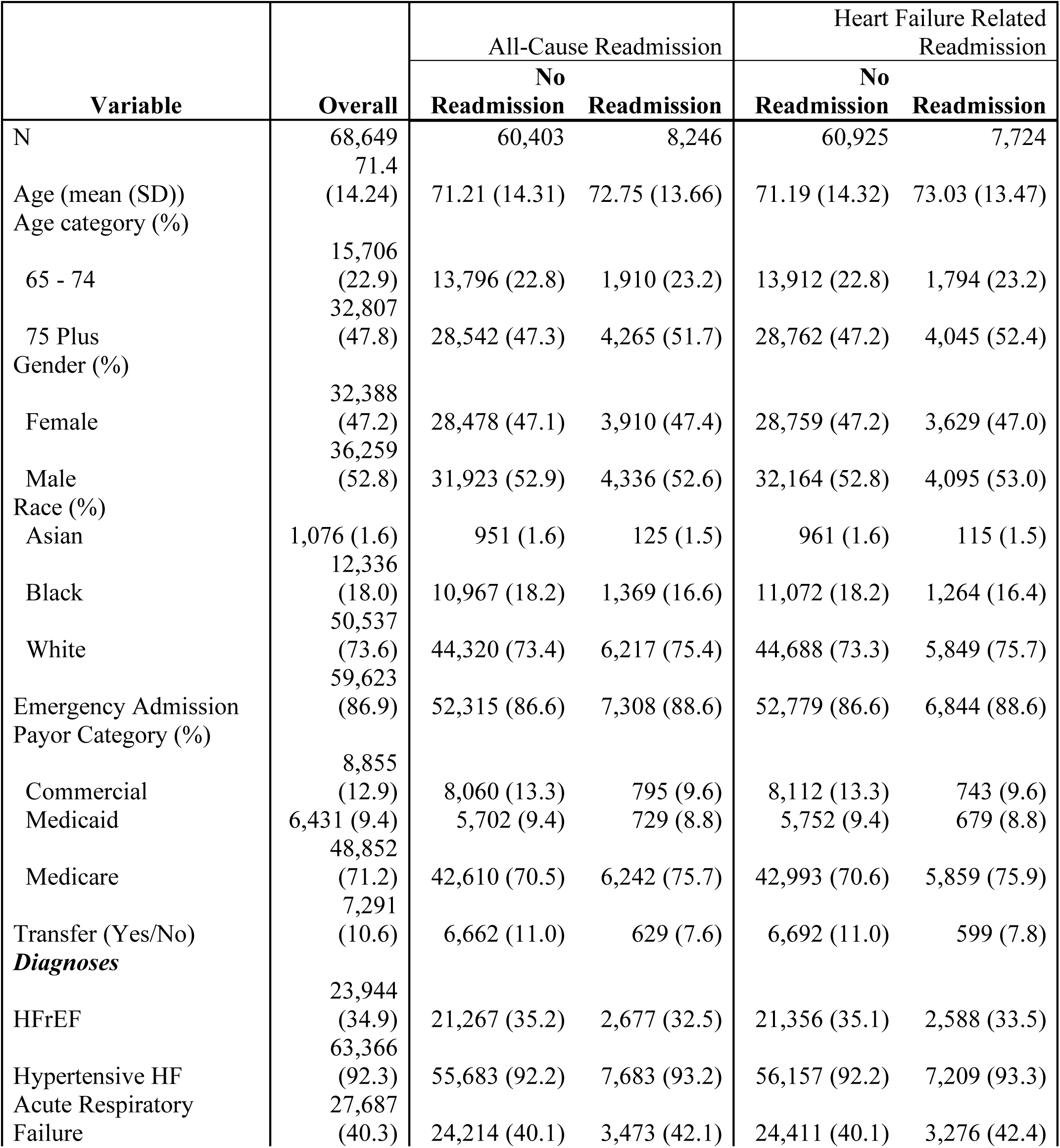
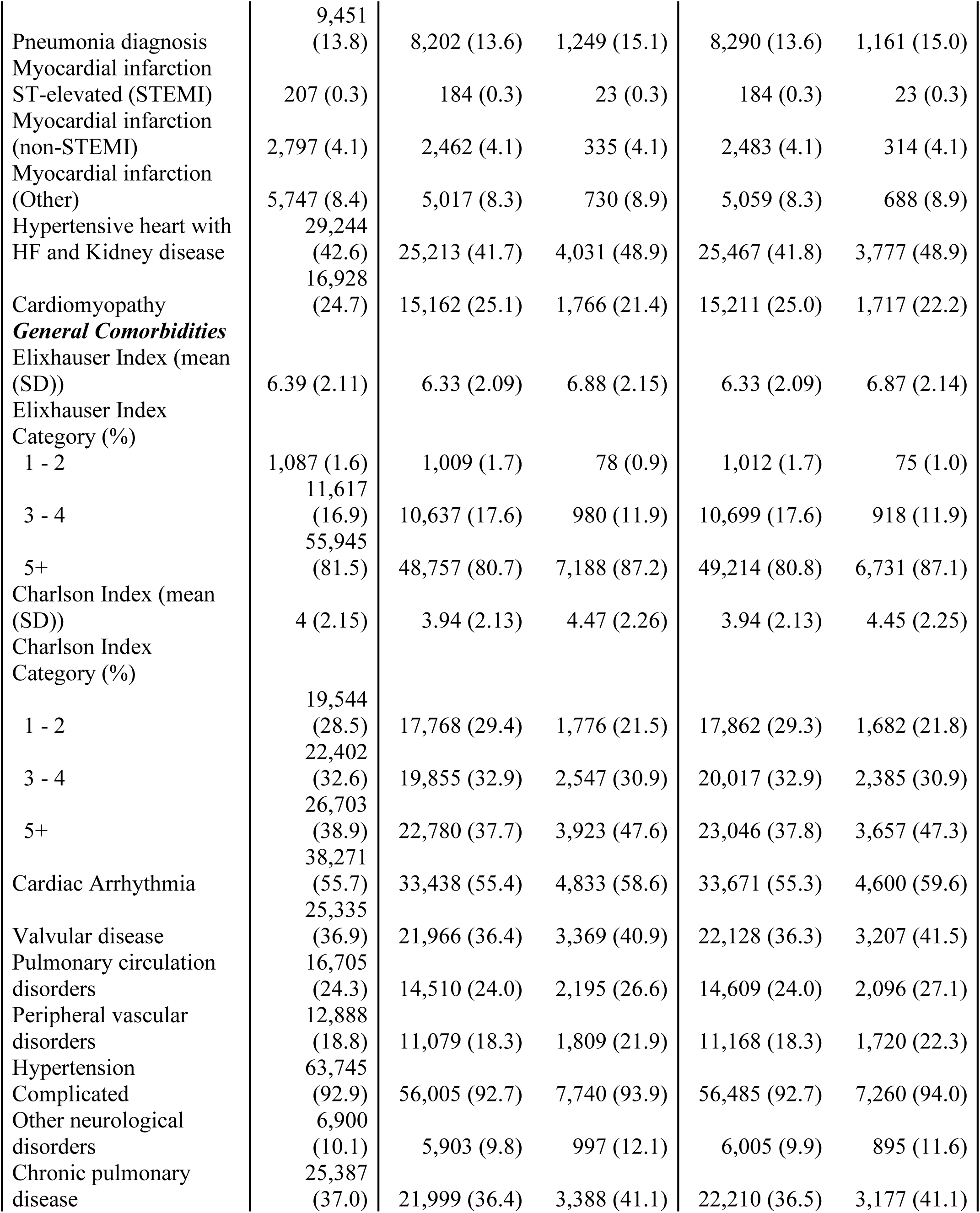
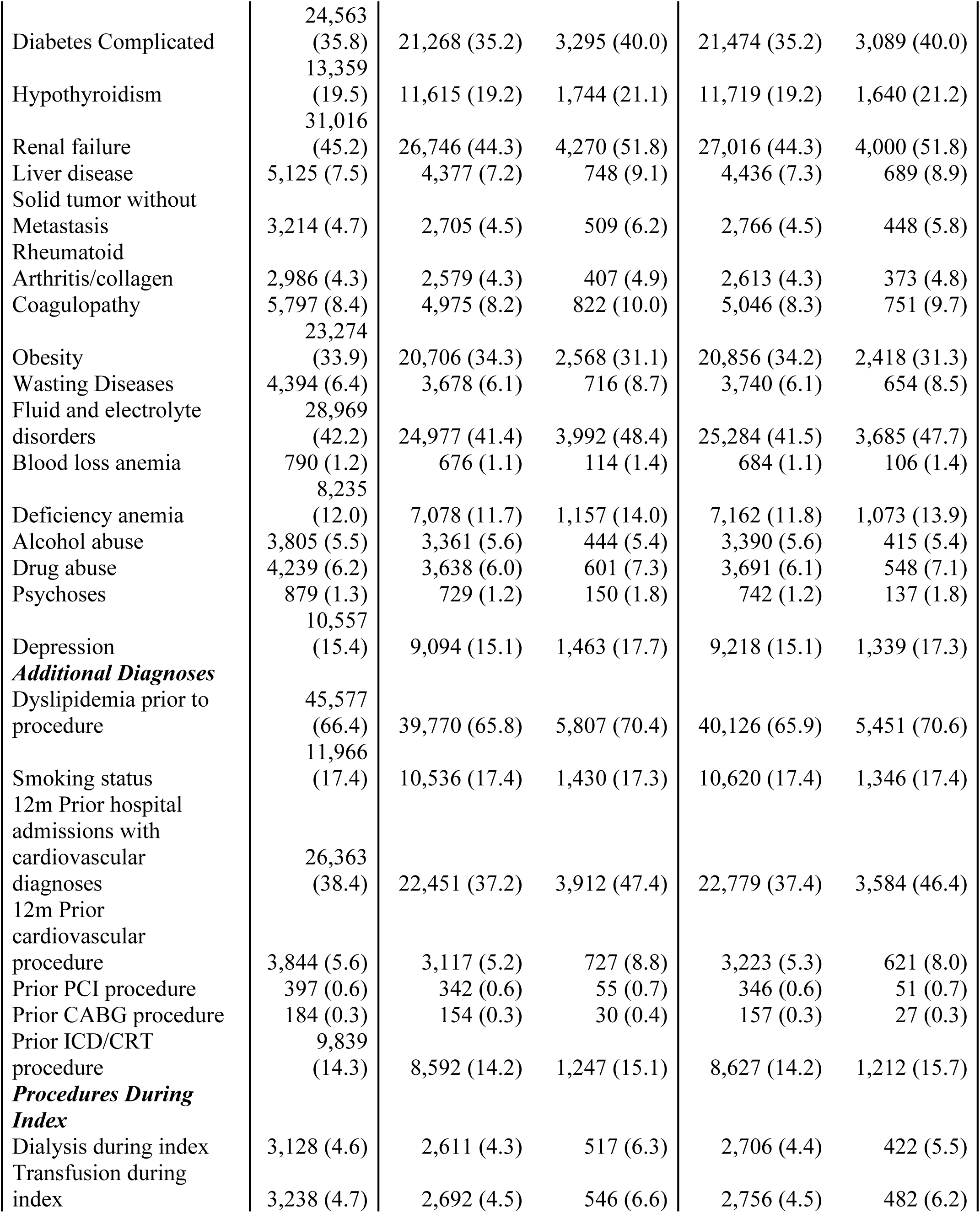
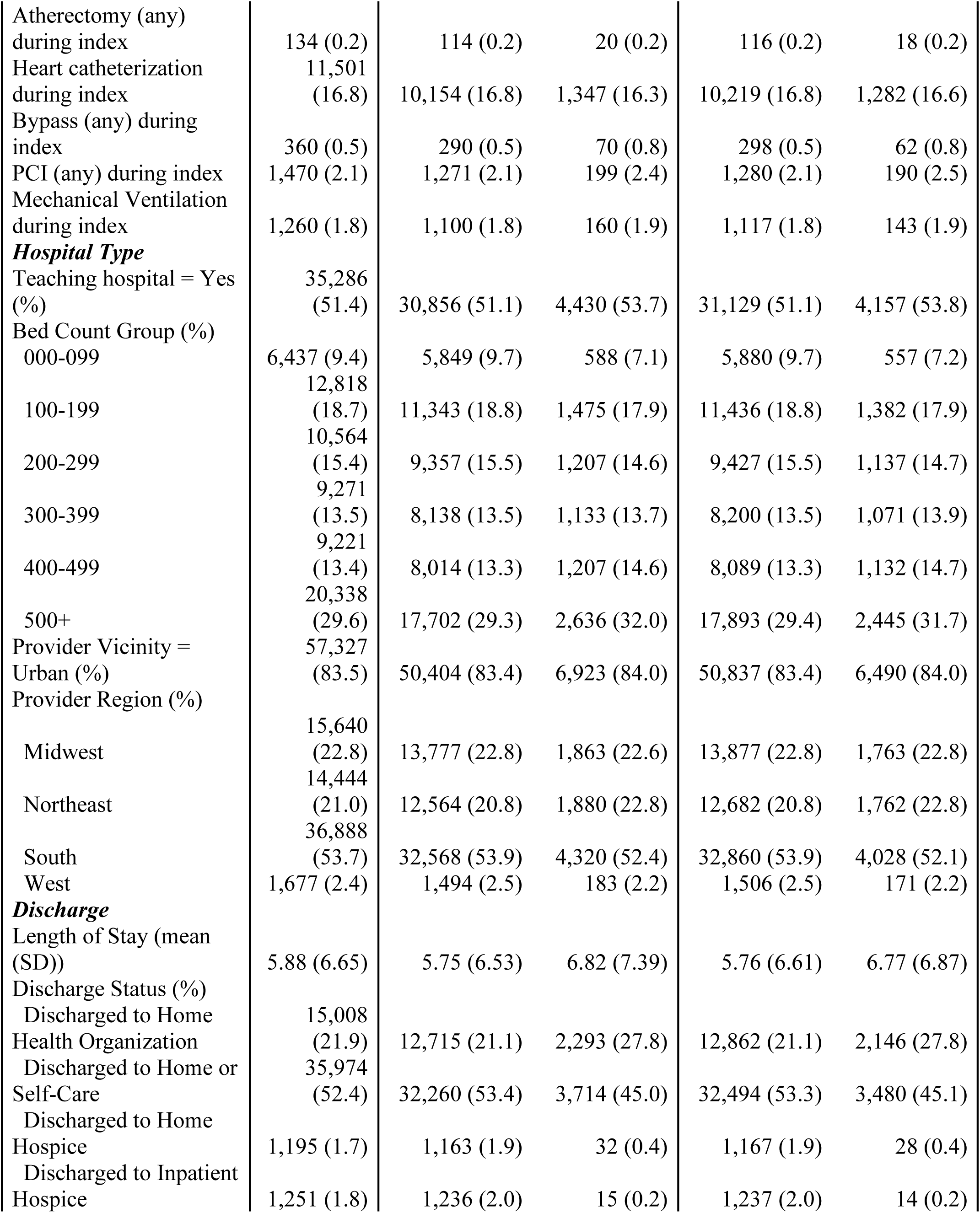
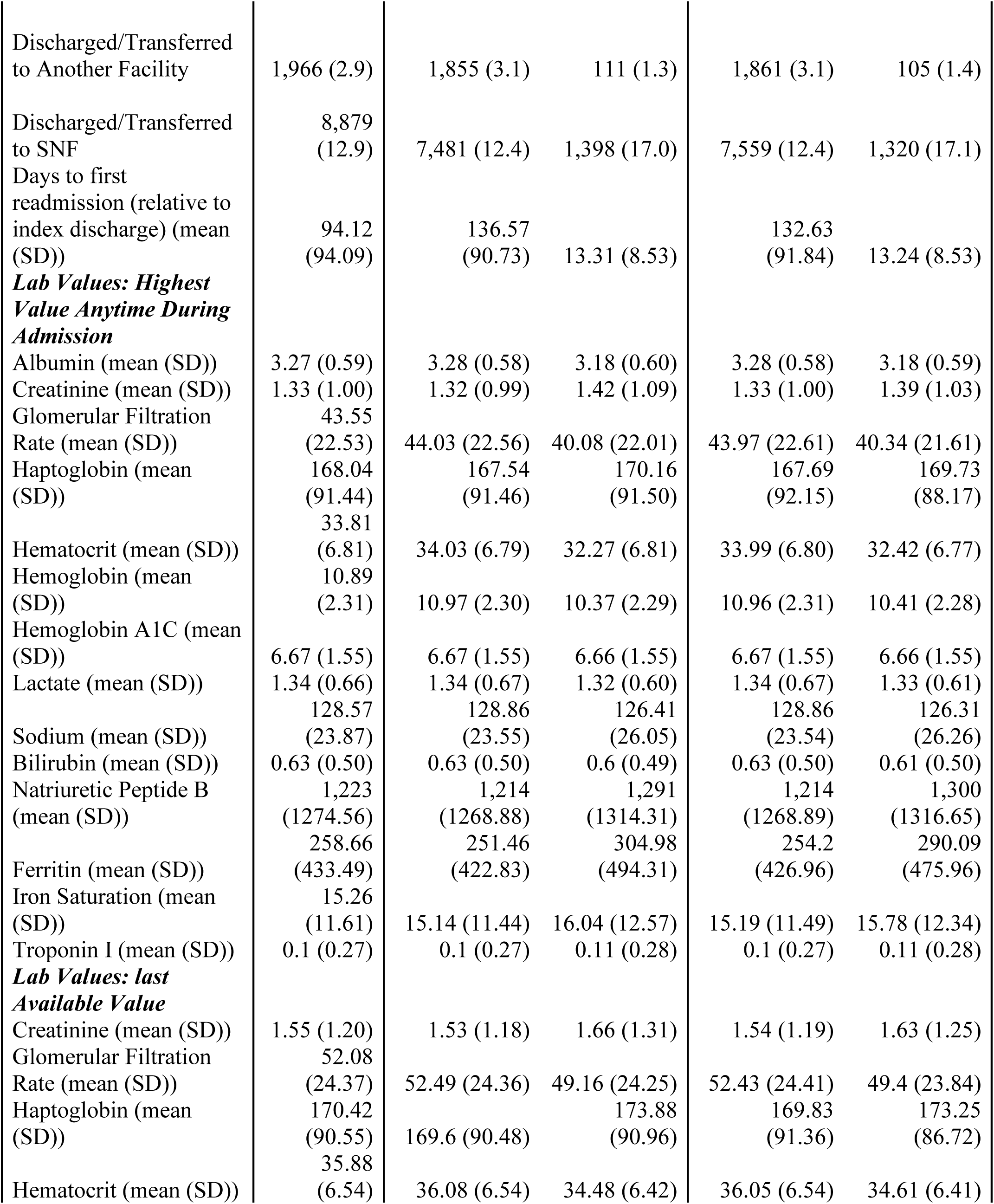
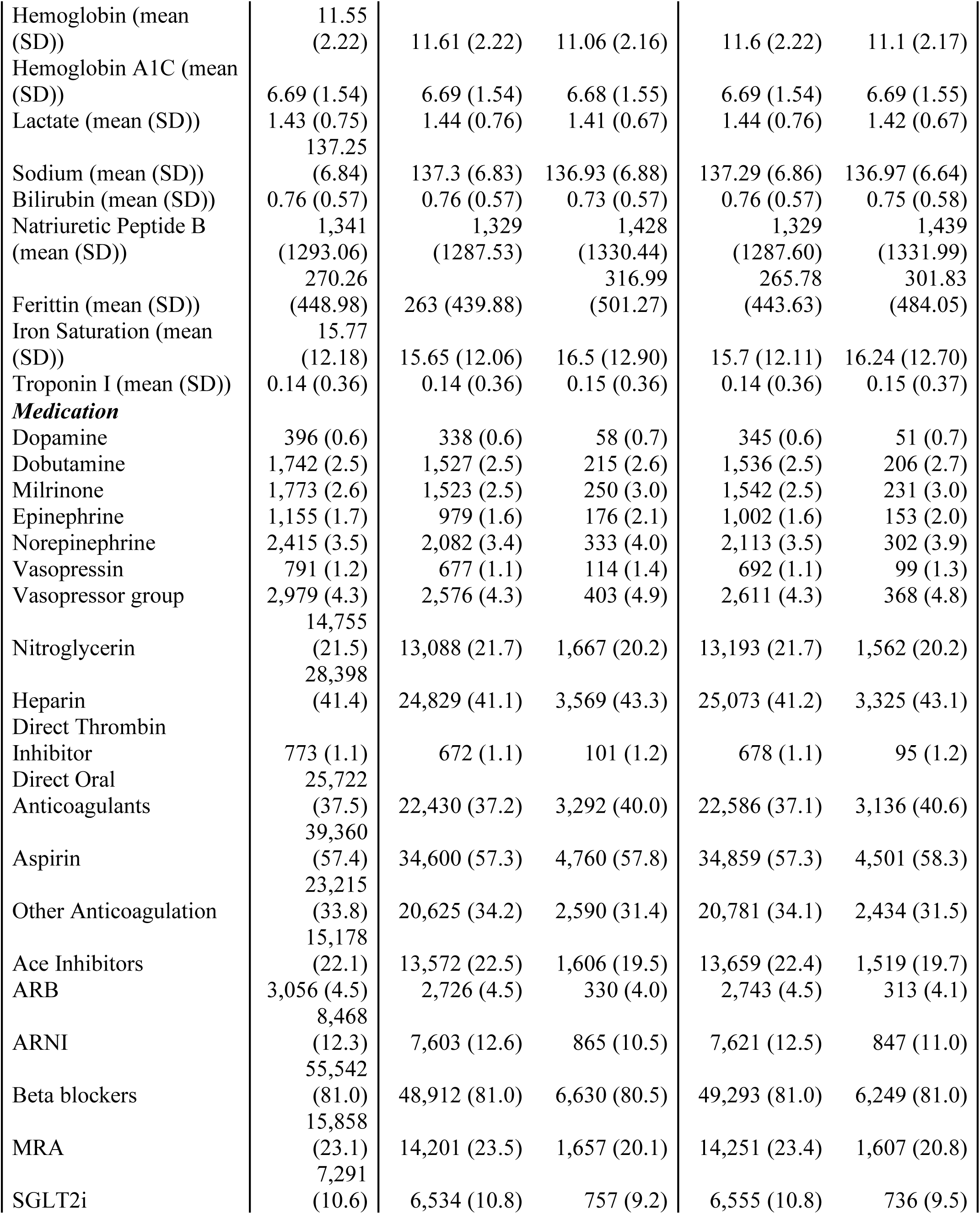
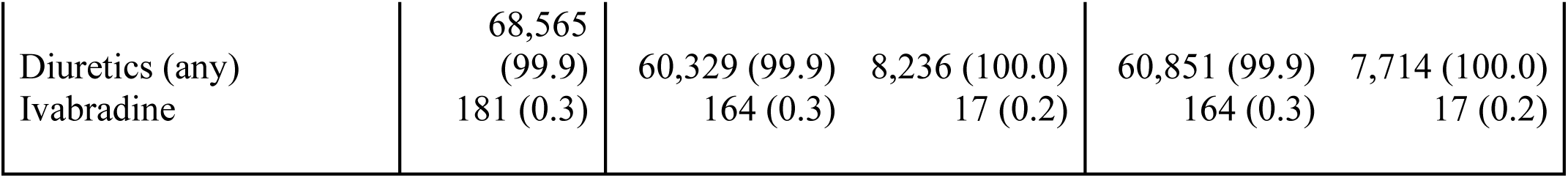
Key Demographic, Clinical and Provider Characteristics of Patients with a Primary or Admitting Diagnosis of Heart Failure 2021-2023.

Two key family of diagnoses were used to identify HF: 1) ICD-10 I50: Heart Failure; and 2) Subsets of ICD-10 I11 and I13 codes (hypertensive heart and chronic kidney disease) that specify presence of heart failure. The COVID-19 era (Q4 2019 to Q1 2021) was excluded, while 2016 through Q3 2019 were categorized as the pre-COVID-19 era and 2021-2023 were categorized as the post-COVID-19 era.

### Variables and Definitions

Since the goal of the study was to predict readmission, patients who died during the index admission were excluded. Only patients from hospitals that provide laboratory values to PHD were included, so that laboratory values could be included in the models. The first inpatient admission with the primary diagnosis of HF for any given patient was defined as the “index,” all subsequent visits were defined as readmissions. Our two outcomes: 1) all-cause 30-day inpatient readmission; and 2) 30-day readmission with a diagnosis (primary, admitting or secondary) of heart failure.

Patient demographics (age, gender, payer, regions, race), comorbidity (defined by presence of chronic diseases, as required to calculate the Elixhauser Comorbidity Index^8^, as well as the actual calculated Elixhauser comorbidity index), and hospital characteristics (size, rural vs urban, teaching status and region) were identified for all patients. In addition, all cardiovascular diagnoses were identified and characterized as present on admission, present in the pre-admission period or present during inpatient care (but not on admission). Medications specific to cardiovascular care were analyzed for all patients, as well as lab values. Medications were either analyzed as individual or grouped based on medication type, and included: ACEi, ARB, ARNI, ARNI, MRA, SGLT2i, diuretics, digoxin, dopamine, dobutamine, milrinone, vasopressor, NTG, and beta blockers. Lab values included albumin, creatinine, glomerular filtration rate, haptoglobin, hematocrit, hemoglobin, HbA1c, lactate, sodium bilirubin, natriuretic peptide B, ferritin, iron saturation, and troponin.

### Statistical Analysis

Descriptive tables with all variables were created (Table 1). The statistical significance of each variable was tested using parametric or non-parametric techniques based on variable type. For parametric variables, t-test for continuous and Pearson’s Chi Square test for categorical variables were used. For non-parametric variables, Mann Whitney tests were used. Collinearity and correlation assessment were used to identify variables for parsimonious modeling.

The data were randomly sampled and partitioned into training (60%), validation (20%) and testing (20%) datasets. The following models were initially tested: Logistic regression, random forest, Decision trees, KNN, Naive Bayes, Support Vector Machines (SVM), neural networks and XGboost. The accuracy of the models was measured using the following metrics: model accuracy (AUC), precision, recall, F-score. Additional models were created using XGboost, as this method provided highest accuracy in preliminary testing. The additional models were based on the following populations: 1) all patients, 2) only patients with primary or admitting (PA) diagnoses of heart failure (excluding those with heart failure diagnosis as secondary diagnosis only), hitherto referred to as the PA Cohort, 3) patients with PA diagnoses in the post-COVID years (Q2 2021 – Q4 2023), and 4) patients with PA diagnoses, in the post-COVID years, with at least one lactate lab value available.

## RESULTS

### Patients

Of 722,974 HF patients identified, a total of 68,649 patients with a PA diagnosis of heart failure in the post-COVID era were identified (Table 1). The average age was 72.1 years, the majority (52.8%) were male, and 74.3% were White. The Elixhauser Index across the entire cohort averaged 6.42 (SD: 2.12), indicating a high level of chronic comorbidity in the population, with 81.7% patients having an Elixhauser Index of 5 or above. At time of index HF admission, 40.6% presented with acute respiratory failure, 13% with pneumonia, 25% with cardiomyopathy and 46.5% with diagnosed atherosclerosis. Key chronic conditions included cardiac arrythmia (57.3%) and renal failure (44.8%). The majority of patients with 30-day readmissions (2,099/2,224, or 94.3%) had a readmission with a HF diagnosis.

### Model Performance

Predictive models were fit for 30-day all-cause and HF specific readmission using 246 candidate variables and eight modeling approaches (Table 2). The models were balanced using a scaling factor and across both outcomes, the XGboost had the best performance. For all-cause readmission the XGboost performance was accompanied by an AUC of 0.625. When modeling 30-day HF specific readmission the model had an AUC of 0.622. We used the XGBoost imputation algorithm, Sparsity-aware split-finding to address any missingness and duly tested for overfitting.

**Table 2.**
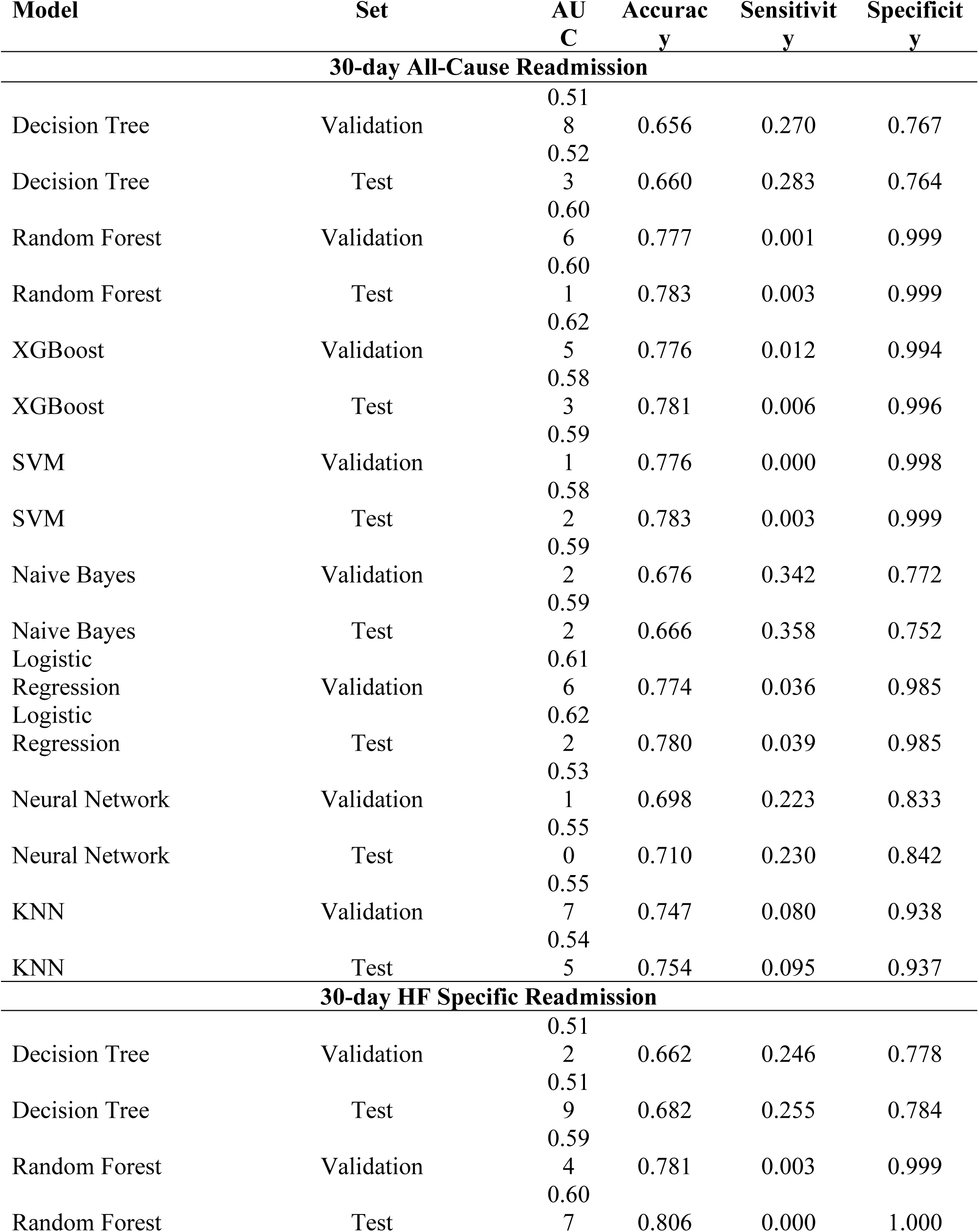
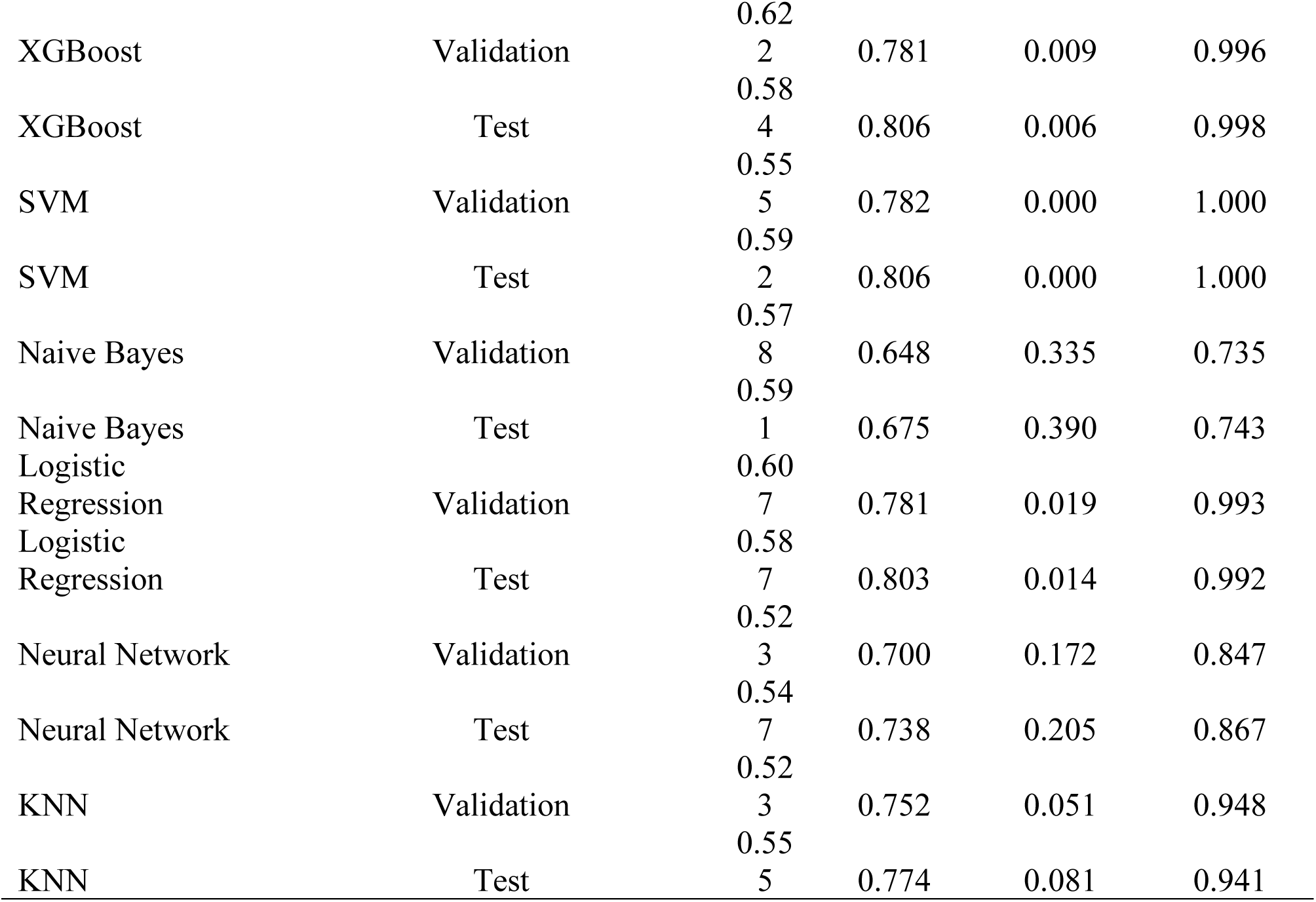
Model Performance.

### Predictors of Readmission

Using the 30-day all-cause readmission XGboost model the most important predictor variables included non-modifieable socioeconomic characteristics and HF related health attributes but not laboratory values or medications (Table 3, Figure A). Similarly, the 30-day HF specific readmission XGboost model highlited non-modifable predictors for readmission (Table 4, Figure B).

**Table 3.** XGBOOST 30-day All-Cause Readmission.

|  |  |  |  |
| --- | --- | --- | --- |
| **Model parameters** |  |  |  |
| colsample_bytree | 0.8 |  |  |
| learning_rate | 0.05 |  |  |
| max_depth | 5 |  |  |
| n_estimators | 100 |  |  |
| sub_sample | 0.8 |  |  |
| **Metrics** | **Train** | **Validation** | **Test** |
| Sample size (N) | 43317 | 10830 | 13537 |
| ROC_AUC | 0.74 | 0.63 | 0.58 |
| F1_Score | 0.33 | 0.26 | 0.25 |
| Recall | 0.66 | 0.53 | 0.53 |
| Precision | 0.22 | 0.17 | 0.17 |
| Accuracy | 0.67 | 0.63 | 0.63 |
| **Top 20 variables (feature importance)** |  |  |  |
| **Feature** | **Importance** |  |  |
| Prior Admission with a Cardiac Diagnosis | 0.0301 |  |  |
| Atherosclerosis Diagnosed in past 12 months | 0.0162 |  |  |
| Elixhauser Category | 0.0143 |  |  |
| Home Discharge | 0.0139 |  |  |
| Length of Stay of Index Admission | 0.0139 |  |  |
| Hemoglobin: Latest Value Before Discharge | 0.0136 |  |  |
| Transfer Required for Index | 0.0121 |  |  |
| Discharge with HHO | 0.0114 |  |  |
| Obesity | 0.0113 |  |  |
| Hematocrit: Latest Value Before Discharge | 0.0113 |  |  |
| Age Group | 0.0109 |  |  |
| Hospital Size (in Bed Count) | 0.0106 |  |  |
| Valvular Disease | 0.0098 |  |  |
| Hypothyroidism | 0.0097 |  |  |
| Chronic pulmonary disease | 0.0096 |  |  |
| Heart Monitoring at Index | 0.0093 |  |  |
| Drug abuse | 0.0090 |  |  |
| Depression | 0.0089 |  |  |
| MRA at Index | 0.0087 |  |  |
| Psychoses | 0.0086 |  |  |

**Table 4.** XGBOOST 30-day HF Specific Readmission.

|  |  |  |  |
| --- | --- | --- | --- |
| **Model parameters** |  |  |  |
| colsample_bytree | 0.8 |  |  |
| learning_rate | 0.05 |  |  |
| max_depth | 5 |  |  |
| n_estimators | 100 |  |  |
| sub_sample | 0.8 |  |  |
| **Metrics** | **Train** | **Validation** | **Test** |
| Sample size (N) | 43317 | 10830 | 13537 |
| ROC_AUC | 0.73 | 0.62 | 0.58 |
| F1_Score | 0.30 | 0.24 | 0.24 |
| Recall | 0.72 | 0.58 | 0.60 |
| Precision | 0.19 | 0.15 | 0.15 |
| Accuracy | 0.62 | 0.59 | 0.57 |
| **Top 20 variables (feature importance)** |  |  |  |
| **Feature** | **Importance** |  |  |
| Prior Admission with a Cardiac Diagnosis | 0.0247 |  |  |
| Elixhauser Category | 0.0211 |  |  |
| Length of Stay of Index Admission | 0.0163 |  |  |
| Discharge with HHO | 0.0137 |  |  |
| Medicare Payer | 0.0134 |  |  |
| Hemoglobin: Latest Value Before Discharge | 0.0121 |  |  |
| Age Group | 0.0104 |  |  |
| Atrial Fibrillation Diagnosed in 12 Months Pre-Index | 0.0102 |  |  |
| Transfer Required for Index | 0.0102 |  |  |
| Obesity | 0.0096 |  |  |
| Valvular Disease | 0.0095 |  |  |
| Dyslipidemia | 0.0093 |  |  |
| Home Discharge | 0.0089 |  |  |
| Hospital Size (in Bed Count) | 0.0088 |  |  |
| Hematocrit: Latest Value Before Discharge | 0.0088 |  |  |
| Chronic pulmonary disease | 0.0086 |  |  |
| Drug abuse | 0.0086 |  |  |
| Admission through non-Healthcare Clinic | 0.0083 |  |  |
| Milrinone Use at Index | 0.0083 |  |  |
| Cardiac Arrhythmia | 0.0083 |  |  |

## DISCUSSION

Our study evaluated over 700,000 HF patients and applied several modeling strategies to predict 30-day readmission in a large nationally representative all-payor dataset. The all-cause readmission rate was 12.0% and the HF-specific readmission rate was 11.3%, lower than previously reported national estimates of approximately 20%.^3^ The AUC values for both all-cause and HF-specific readmission were 0.63 and 0.62, respectively, indicative that, despite contemporary predictive modeling approaches, HF readmission may not be attributed to modifiable risk factors primarily. Key predictors of readmission may be therefore largely non-modifiable, without explicit actionable interventions. These included age, socioeconomic status, and other comorbidities rather than HF-specific therapies or laboratory values. These results underscore the ongoing challenge in predicting and preventing HF readmissions and question the rationale and efficacy of federal fiscal penalties in the quest for reductions in readmission rates.

Predicting and preventing HF readmissions remains an inherently complex challenge, as demonstrated by the poor performance of previous models.^9–11^ Management of HF encompasses highly individualized treatment that is often influenced by medical, behavioral, and socioeconomic factors that are difficult to quantify in traditional claims-based data models.

Mitigation strategies include the expansion of post-discharge telephone or telemedicine connectivity and home-based monitoring programs, often incorporating multidisciplinary teams.^12–16^ Emerging solutions may involve real-time patient monitoring using wearable sensors, artificial intelligence-driven risk stratification tools, and personalized HF management programs. Additionally, greater integration of mental health and social support services into HF care may help address non-clinical drivers of readmission risk, such as medication adherence and access to follow-up care.^17,18^

The Hospital Readmissions Reduction Program (HRRP) was introduced under the Affordable Care Act (ACA) to financially penalize hospitals with higher-than-expected readmission rates, with the goal of improving patient outcomes and reducing unnecessary healthcare expenditure.^4^ While this policy has admittedly led to some reductions in national readmission rates, evidence remains mixed regarding its long-term efficacy and equity.^3,4,18,19^ Studies have demonstrated that HRRP may disproportionately disadvantage safety-net hospitals and those that serve socioeconomically disadvantaged populations, leading to potential reductions in hospital reimbursement without commensurate improvements in patient care.^21,22^ Though beneficial, the 2019 implementation of social risk stratification still is not ubiquitous across the landscape of healthcare systems caring for the most vulnerable.^23,24^ Furthermore, the focus on 30-day readmission rate as a key quality metric may not adequately capture the complexity of HF management, as many readmissions are driven by non-modifiable patient factors rather than deficiencies in care quality.^4,7,21^ The potential unintended consequence of financially incentivizing reduction of readmissions may potentially disincentivize the care of higher-risk patients.^4,19,20^ Instead, perhaps the HRRP could be redesigned to follow risk-adjusted quality metrics that guide the adjudication of readmission penalties and account for non-modifiable factors common to these patients such as cumulative non-cardiac comorbidities, social determinants of health, geography, and healthcare access.^4^

The study has several limitations. First, as a retrospective analysis based on a large administrative dataset, the findings are subject to unmeasured confounders. Additionally, while the dataset captures a broad representation of hospitals, it excludes Veterans Affairs hospitals, potentially limiting generalizability. The exclusion of the COVID-19 period may impact findings, given that healthcare delivery underwent significant unexpected changes although by doing this we sought to eliminate pandemic-associated bias.^25^ Despite these clear temporal and data limitations, this study attempted to compensate by examining a large nationally-representative all-payor dataset by deploying advanced machine learning methodology and, unlike conventional logistic regression models that assume linearity, the use of XGBoost and other machine learning models may permit more complex interactions between variables.^9^

In conclusion, federal programs that seek to reduce readmissions through financial penalties by focusing on modifiable HF-specific factors may not optimally achieve an improvement in patient care, as it appears the primary driver of HF-readmission may be non-modifiable factors. Our findings suggest that these penalties may be punitive but without corrective or actionable counter interventions. Given the predominance of non-modifiable nature of the risk factors in HF readmissions, future policy adjustments should incorporate broader measures of care quality and risk-adjusted patient-centered outcomes. Further research is needed to refine predictive modeling techniques, integrate real-world patient data, and develop targeted interventions that address both medical and socioeconomic determinants of readmission.

## Data Availability

There is no additional data referred to within the text that is not included.

## Acknowledgments

The authors acknowledge and thank Chantal Holy, MSc PhD for her statistical support.

---

**Figure A.**
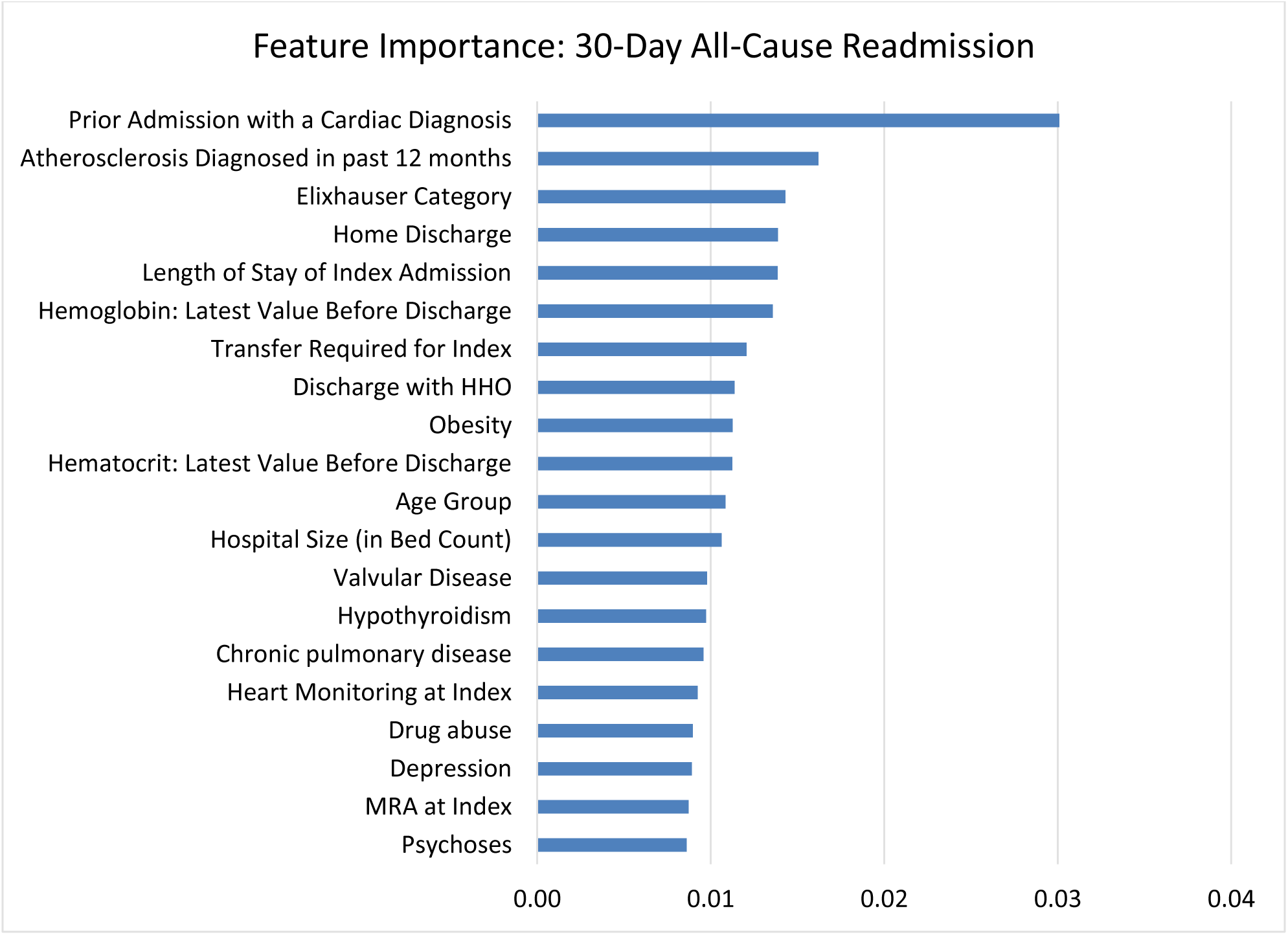
Feature Importance: 30-Day All-Cause Readmission. Listed are the top 20 features of importance portending all-cause readmission within 30 days.

**Figure B.**
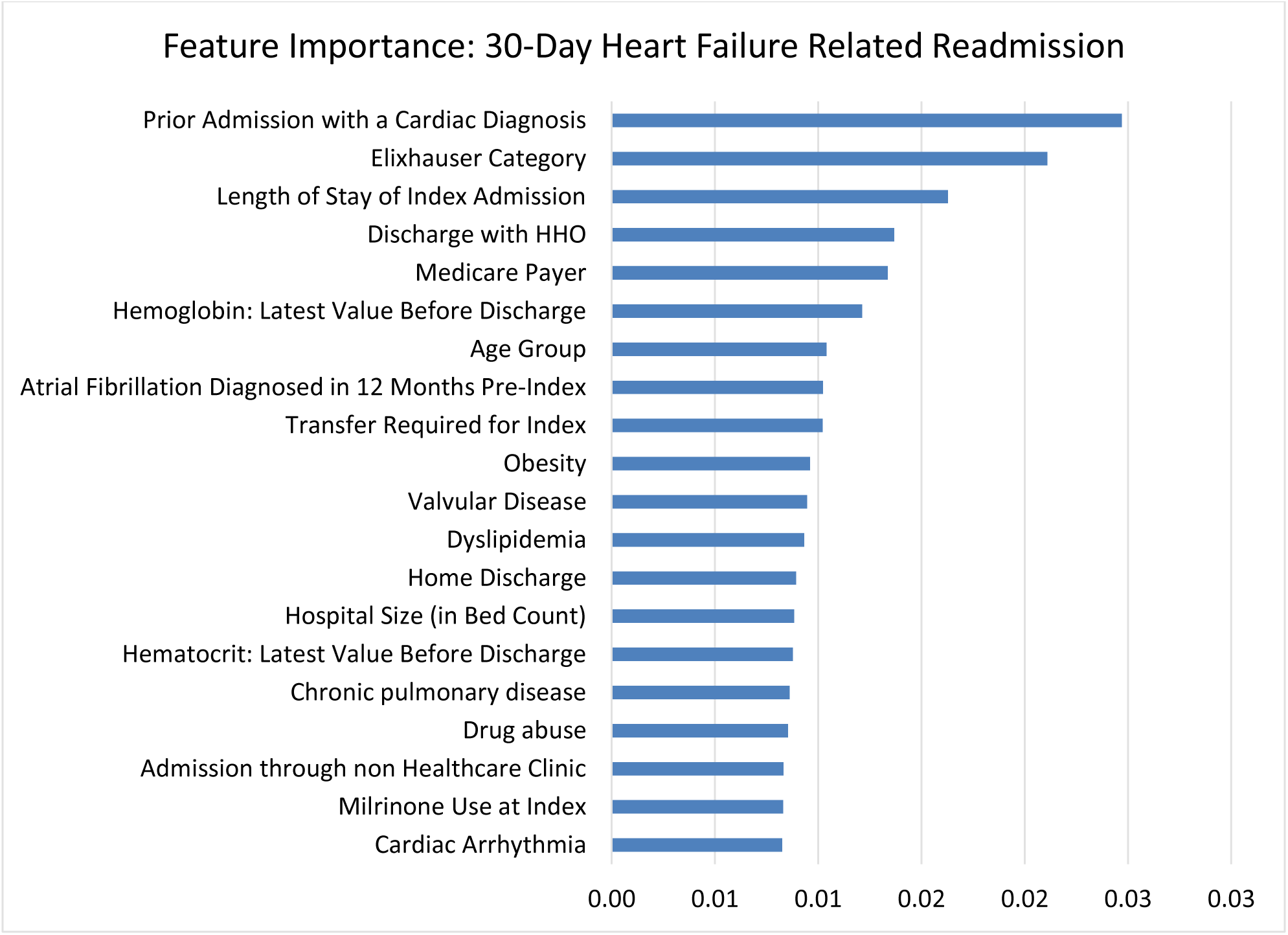
Feature Importance: 30-Day Heart Failure Related Readmission. Listed are the top 20 features of importance portending readmission for heart failure within 30 days.

